# Late immune-related adverse events in long-term responders to PD-1/PD-L1 checkpoint inhibitors: a multicentre study

**DOI:** 10.1101/2020.04.09.20054031

**Authors:** Olga Nigro, Graziella Pinotti, Federica De Galitiis, Francesca Romana Di Pietro, Raffaele Giusti, Marco Filetti, Melissa Bersanelli, Alessandro Lazzarin, Paola Bordi, Annamaria Catino, Pamela Pizzutilo, Domenico Galetta, Paolo Marchetti, Andrea Botticelli, Simone Scagnoli, Marco Russano, Daniele Santini, Mariangela Torniai, Rossana Berardi, Biagio Ricciuti, Andrea De Giglio, Rita Chiari, Alessandro Russo, Vincenzo Adamo, Marianna Tudini, Rosa Rita Silva, Elena Bolzacchini, Monica Giordano, Pietro Di Marino, Michele De Tursi, Erika Rijavec, Michele Ghidini, Ilaria Vallini, Luigia Stefania Stucci, Marco Tucci, Laura Pala, Fabio Conforti, Paola Queirolo, Enrica Tanda, Francesco Spagnolo, Federica Cecchi, Sergio Bracarda, Serena Macrini, Matteo Santoni, Nicola Battelli, Maria Concetta Fargnoli, Giampiero Porzio, Alessandro Tuzi, Matteo Basilio Suter, Corrado Ficorella, Alessio Cortellini

## Abstract

Data on spectrum and grade of immune-related adverse events (irAEs) in long-term responders to immune checkpoint inhibitors (ICI) are lacking.

We performed a retrospective multicenter study to characterized irAEs occurring after a 12-months minimum treatment period with PD-(L)1 ICIs in advanced cancer patients. IrAEs were categorized into “early” (≤12 months) and “late” (>12 months).

From September 2013 to October 2019, 436 consecutive patients were evaluated. 223 experienced any grade early-irAEs (51.1%), while 132 experienced any grade late-irAEs (30.3%) (p < 0.0001). Among the latter, 29 (22%) experienced a recurrence of an early-irAEs, while 103 (78%) experienced *de novo* late-irAEs involving different system/organ. Among patients with late-irAEs, 21 experienced G3/G4 irAEs (4.8%). Median time to onset of early-irAEs was 3.4 months (95%CI: 2.8-4.2), while the median time to onset of late-irAEs was 16.6 months (95%CI: 15.8-17.6). Cumulative time-adjusted risk of disease progression according to both the early-irAEs (HR = 0.63 [95%CI: 0.30-1.29], p = 0.204) and late-irAEs occurrence revealed no statistically significant differences (HR = 0.75 [95%CI: 0.37-1.56], p = 0.452). Also the time-adjusted cumulative risk of death according to both early-irAEs (HR = 0.79 [95%CI: 0.34-1.86], p = 0.598) and late-irAEs (HR = 0.92 [95%CI: 0.49-1.74], p = 0.811) did not show statistically significant differences.

Although less frequent than early-irAEs, late-irAEs are quite common in long responders to PD-(L)1 ICIs, and are different in terms of spectrum and grade. Time-adjusted analysis revealed that the cumulative risk of disease progression and death were not significantly reduced in patients who experienced late-irAEs.

## Introduction

Methods of adverse events (AEs) reporting in clinical trials are aimed at quantifying and qualifying development of AEs over time, which have traditionally been reported as cumulative incidence, highlighting the maximum AE grade per patient during the study period [1]. The greatest limitation of these methods is that they do not consider the duration of treatment, as in exposure-adjusted safety analyses [2]. Tables focusing on high-grade AEs do not depict the evolution of toxicity over time [1]. “Timing” is important for oncological therapies with chronic administration [2, 3], as for Immune Checkpoint Inhibitors (ICIs), which can be associated with immune-related adverse events (irAEs). Updates of prospective studies of anti-programmed death-1 (PD-1) agents demonstrated that safety profile of these drugs remains almost unchanged over time [4-6]. Furthermore, guidelines report that irAEs occur quite early, mostly within weeks from immunotherapy start, and only occasionally after the first year of treatment [7, 8]. Interestingly, studies conducted across different tumor types have shown that irAEs are associated with improved clinical outcomes, and their occurrence has increasingly been interpreted as a pharmacodynamic effect of ICIs [9, 10]. These studies showed conflicting results, especially due to lead-in time bias of time-dependent nature of irAEs [11]. Against this background, we conducted a multicentre, retrospective study on a long-term responder advanced cancer patients, and characterized those irAEs occurring after the first year of immunotherapy.

## Materials and Methods

This multicenter retrospective observational study evaluated advanced cancer patients consecutively treated with single agent PD-1/PD-L1 (programmed death-ligand 1) checkpoint inhibitors (standard schedules) from September 2013 to October 2019, regardless of treatment line, at 20 Italian institutions (Supplementary file 1). Patients were eligible if they had a minimum treatment duration of 12 months.

### Study design

Primary end-point was to describe incidence and characteristics of irAEs occurring after the first 12 months of treatment with PD-1/PD-L1 checkpoint inhibitors (late-irAEs); secondary end-point was to further investigate the relationships between irAEs occurrence, progression free survival (PFS), and overall survival (OS).

IrAEs were categorized into: “early-irAEs” (≤12 months of immunotherapy), and “late-irAEs” (>12 months). IrAEs that started ≤12 months, but lasted more than 12 months, were considered early-irAEs. IrAEs were graduated according to the National Cancer Institute Common Toxicity Criteria for Adverse Events (CTCAE; version 4.0). IrAEs were categorized on the basis of the organ/system involved: cutaneous, endocrine (including thyroid disorders), gastro-intestinal (GI), pneumological, hepatic, rheumatologic, neurologic, and others irAEs (including hyperpyrexia, pancreatitis and asthenia). IrAEs were defined “single-site” if involving one system/organ; “multiple-site” if involving more than one system/organ. Analyses by categories and number of involved sites were performed only for any grade irAEs and not for G3/G4 irAEs. χ2 test was used to evaluate the difference between early- and late-irAEs [12]. Clinical management of early- and late-irAEs was categorized as follows: no intervention (supportive treatments), corticosteroids administration without immunotherapy interruption, corticosteroids administration with temporary interruption, and corticosteroids administration with permanent discontinuation. Late-irAEs which represent a recurrence of an early-irAEs was also described and reported as late-irAEs, when spaced by an irAEs free interval. Median time to onset of early- and late-irAEs since treatment start was also reported.

Treatment duration was defined as “time from treatment’s start to permanent discontinuation” for any reason (disease progression, treatment toxicity, patient preference, death). Baseline patients’ features were reported: age (< 70 vs. ≥ 70 years old) [13-16], sex (male vs female), primary tumor (NSCLC, melanoma, renal cell carcinoma and others), Eastern Cooperative Oncology Group Performance Status (ECOG-PS) (0-1 vs ≥ 2), number of involved organs including the primitive tumor (≥ 3 vs < 3) and treatment line (1st vs ≥ 2nd).

Patient characteristics and objective response rate (ORR) were reported with descriptive statistic. Patients were assessed with radiological imaging every 8-12 weeks according to national guidelines required by Agenzia Italiana del Farmaco (AIFA); RECIST (v. 1.1) criteria were used [17], but treatment beyond disease progression was allowed when clinically indicated. ORR was defined as portion of patients experiencing an objective response (complete or partial response) as best response to immunotherapy; PFS as time from treatment’s start to disease progression or death whichever occurred first; OS as time from the beginning of treatment to death.

Median treatment duration, median time to irAEs onset, median PFS and median OS were evaluated using the Kaplan-Meier method [18]. Median period of follow-up was computed according to the reverse Kaplan-Meier method [19]. Cox proportional hazards model was used to evaluate PFS and OS according to early-irAEs occurrence (yes vs no) and late-irAEs occurrence (yes vs no) and to estimate hazard ratios (HRs) for disease progression and death [20]. Considering the lead-in time bias caused by time-dependent nature of irAEs a further analysis was performed, in order to estimate the risk of disease progression and death over time, according to the early-irAEs and late-irAEs occurrence. A Cox model was therefore fitted by considering irAEs occurrence a time-varying covariate that had a value of 0 before the irAE onset and 1 after irAE onset; cumulative hazard plots was also presented [21]. Data cut-off period was October 2019. Statistical analyses were performed using MedCalc Statistical Software version 18.11.3 (MedCalc Software bvba, Ostend, Belgium; https://www.medcalc.org; 2019). Time-adjusted Cox regression was performed using the R package rpart (R version 3.6.2).

## Results

### Patient characteristics

436 consecutive patients were evaluated; median age was 68 years (range: 32-92). 291 (66.7%) were male, while 145 (33.3%) were female. Primary tumors were: NSCLC (49.1%), melanoma (39.0%), kidney (9.4%), and others (2.5%). Clinico-pathologic characteristics of the entire cohort of patients are summarized in Table 1.

**Table 1:** Patients features.

| Patients | 436 - N (%) |
| --- | --- |
| **Age (years)** |  |
| Median (range) | 68 (32-92) |
| Non-elderly | 243 (55.7) |
| Elderly ($\geq 70$ y.o.) | 193 (44.3) |
| **Gender** |  |
| Male | 291 (66.7) |
| Female | 145(33.3) |
| **Primary Tumor** |  |
| NSCLC | 214 (49.1) |
| Melanoma | 170 (39.0) |
| Kidney | 41 (9.4) |
| Others | 11 (2.5) |
| **ECOG PS** |  |
| 0/1 | 413 (94.7) |
| $\geq 2$ | 23 (5.3) |
| **Number of involved organs** |  |
| $< 3$ | 286 (65.6) |
| $\geq 3$ | 150 (34.4) |
| **Treatment line** |  |
| First | 192 (44.1) |
| Non-first | 244 (55.9) |
| **Immunotherapeutic agents** |  |
| Pembrolizumab | 174 (39.9) |
| Nivolumab | 252 (57.8) |
| Atezolizumab | 10 (2.3) |

### Immune-related adverse events analysis

223 patients experienced any grade early-irAEs (51.1%), while 132 experienced any grade late-irAEs (30.3%), with a statistically significant difference (p<0.0001). Among the latter, 29 (22%) experienced a recurrence of an early-irAEs, while 103 (78%) experienced late-irAEs involving different system/organ not previously involved. 21 patients experienced G3 early-irAEs (4.8%), as well as G3 late-irAEs (21 patients, 4.8%). No G4 early-irAEs, nor G4 late-irAEs were reported. Median time to onset of early-irAEs was 3.4 months (95%CI: 2.8-4.2), while median time to onset of late-irAEs was 16.6 months (95%CI: 15.8-17.6).

Table 2A summarizes early- and late-irAEs according to system/organ involved. Incidence of late cutaneous, endocrine, GI, and other irAEs, was significantly lower compared to incidence of the respective early-irAEs category.

**Table 2:** (A) Any grade early-irAEs and late-irAEs. (B) Clinical management of early-irAEs and late-irAEs. (C) Any grade early-irAEs and late-irAEs in NSCLC and melanoma patients.

**A**
| Any grade irAEs | Early-irAEs (patients-%) | Late-irAEs (patients-%) | P value |
| --- | --- | --- | --- |
| **Overall population** | **223 (51.1)** | **132 (30.3)** | **< 0.0001** |
| Cutaneous | 84 (19.3) | 50 (11.5) | 0.0062 |
| Endocrine | 61 (13.9) | 20 (4.6) | < 0.0001 |
| Gastro-intestinal | 47 (10.8) | 18 (4.1) | 0.0005 |
| Pneumological | 15 (3.4) | 10 (2.3) | 0.3244 |
| Haepatic | 9 (2.1) | 5 (1.1) | 0.2892 |
| Rheumatologic | 43 (9.9) | 29 (6.7) | 0.1129 |
| Neurologic | 1 (0.2) | 4 (0.9) | 0.3735 |
| Others | 52 (11.9) | 13 (2.9) | < 0.0001 |
| **Single-site irAEs** | **149 (34.2)** | **114 (26.1)** | **0.0585** |
| **Multiple-site irAEs** | **74 (17)** | **18 (4.1)** | **< 0.0001** |

**B**
| IrAEs Management | Early-irAEs | Late-irAEs | P value |
| --- | --- | --- | --- |
| No intervention (only supportive) | 110 (49.3) | 59 (44.7) | 0.7789 |
| Corticosteroids without discontinuation | 79 (35.4) | 43 (32.5) | 0.8549 |
| Corticosteroids with temporary interruption | 34 (15.2) | 15 (11.4) | 0.4463 |
| Corticosteroids with permanent discontinuation | - | 15 (11.4) | 0.0001 |

**C**
| Any grade late-irAEs | Melanoma (%) | NSCLC (%) |
| --- | --- | --- |
| **Overall population** | **49 (28.8)** | **77 (35.9)** |
| Cutaneous | 17 (10) | 31 (14.5) |
| Endocrine | 7 (4.1) | 13 (6.1) |
| Gastro-intestinal | 6 (3.5) | 12 (5.6) |
| Pneumological | 3 (1.8) | 5 (2.3) |
| Haepatic | 2 (1.2) | 3 (1.4) |
| Rheumatologic | 7 (4.1) | 20 (9.3) |
| Neurologic | 4 (2.4) | - |
| Others | 10 (5.9) | 2 (0.9) |
| **Single-site irAEs** | **42 (24.7)** | **67 (31.3)** |
| **Multiple-site irAEs** | **7 (4.1)** | **10 (4.7)** |

149 patients in early-irAEs group (34.2%) experienced single-site irAEs, compared to 114 patients (26.1%) in late-irAEs group, (p=0.0585). Similarly, 74 patients in early-irAEs group experienced multiple-site irAEs (17%), compared 18 patients (4.1%) in late-irAEs group, (p<0.0001). Table 2B reports the clinical management of early- and late-irAEs. Early irAEs were managed with supportive interventions only (49.3%), and to a lesser extent with corticosteroids administration with (15.2%) or without (35.4%) temporary immunotherapy discontinuation. Late-irAEs were also managed with supportive interventions only (44.7%) or with corticosteroids administration without immunotherapy discontinuation (32.5%); 11.4% of late-irAEs were managed with corticosteroids administration and temporary discontinuation, as well as corticosteroids administration with permanent discontinuation.

### Clinical outcomes analysis

ORR to immunotherapy in the entire cohort of patients was 71.3% (95%CI: 63.6–79.7): 119 stable disease (27.3%), 250 partial responses (57.3%), 61 complete responses (14.0%), and 6 (1.4%) progressive disease were reported. At a median follow-up of 23.3 months (95%CI: 21.8-25.2), median treatment duration was 30.8 months (95%CI: 27.8-33.6) and 273 patients were still receiving immunotherapy at the data cut-off.

In the overall population, median PFS was 43.1 months (95%CI: 36.6-53.5; 124 events), while median OS was not reached (373 censored). Table 3 summarized conventional Cox proportional hazard estimation for PFS and OS, and time-adjusted cumulative hazard estimation of disease progression and death were presented according to early- and late-irAEs occurrence. Among patients who experienced and who did not experience any grade early-irAEs median PFS was 53.5 months (95%CI: 41.1-53.5; 61 events) and 42.7 moths (95%CI: 31.1-42.7; 63 events), respectively, with no statistically significant difference (HR=0.43 [95%CI: 0.61-1.23], p=0.8702) (Figure 1A). Median PFS among patients who experienced and who did not experience any grade late-irAEs was not reached (27 events) and 41.1 months (95%CI: 30.2-53.5; 97 events), respectively, with a statistically significant difference (HR=0.46 [95%CI: 0.30-0.72], p=0.0005) (Figure 1B). In both the cohorts of patients who experienced (190 censored) and who did not experience any grade early-irAEs (183 censored), median OS was not reached, without statistically significant difference (HR=0.96 [95%CI: 0.58-1.57], p=0.8750) (Figure 1C). Similarly, in both the cohorts of patients who experienced (120 censored) and who did not experience any grade early-irAEs (253 censored), median OS was not reached, but with a statistically significant difference (HR=0.41 [95%CI: 0.22-0.77], p=0.0053) (Figure 1D).

**Table 3:** Subgroup analysis of PFS and OS according to the early-irAEs and late-irAEs occurrence. Conventional Cox proportional hazard estimation and time-adjusted cumulative hazard estimation of disease progression and death were presented.

|  | Conventional Cox hazard estimation of Disease Progression | Conventional Cox hazard estimation of Death |
| --- | --- | --- |
| VARIABLE | HR (95% CI); *p value* | HR (95% CI); *p value* |
| **Early-irAEs**<br>Yes vs No | 0.43 (0.61–1.23); *p=0.8702* | 0.96 (0. 58–1.57); *p=0.8750* |
| **Late-irAEs**<br>Yes vs No | 0.46 (0.30–0.72); *p=0.0005* | 0.41 (0.22–0.77); *p=0.0053* |
|  | Time-adjusted Cox hazard estimation of Disease Progression | Time-adjusted Cox hazard estimation of Death |
| **Early-irAEs**<br>Yes vs No | 0.63 (0.30–1.29); *p=0.204* | 0.79 (0.34–1.86); *p=0.598* |
| **Late-irAEs**<br>Yes vs No | 0.75 (0.37–1.56); *p=0.452* | 0.92 (0.49–1.74); *p=0.811* |

**Figure 1:**
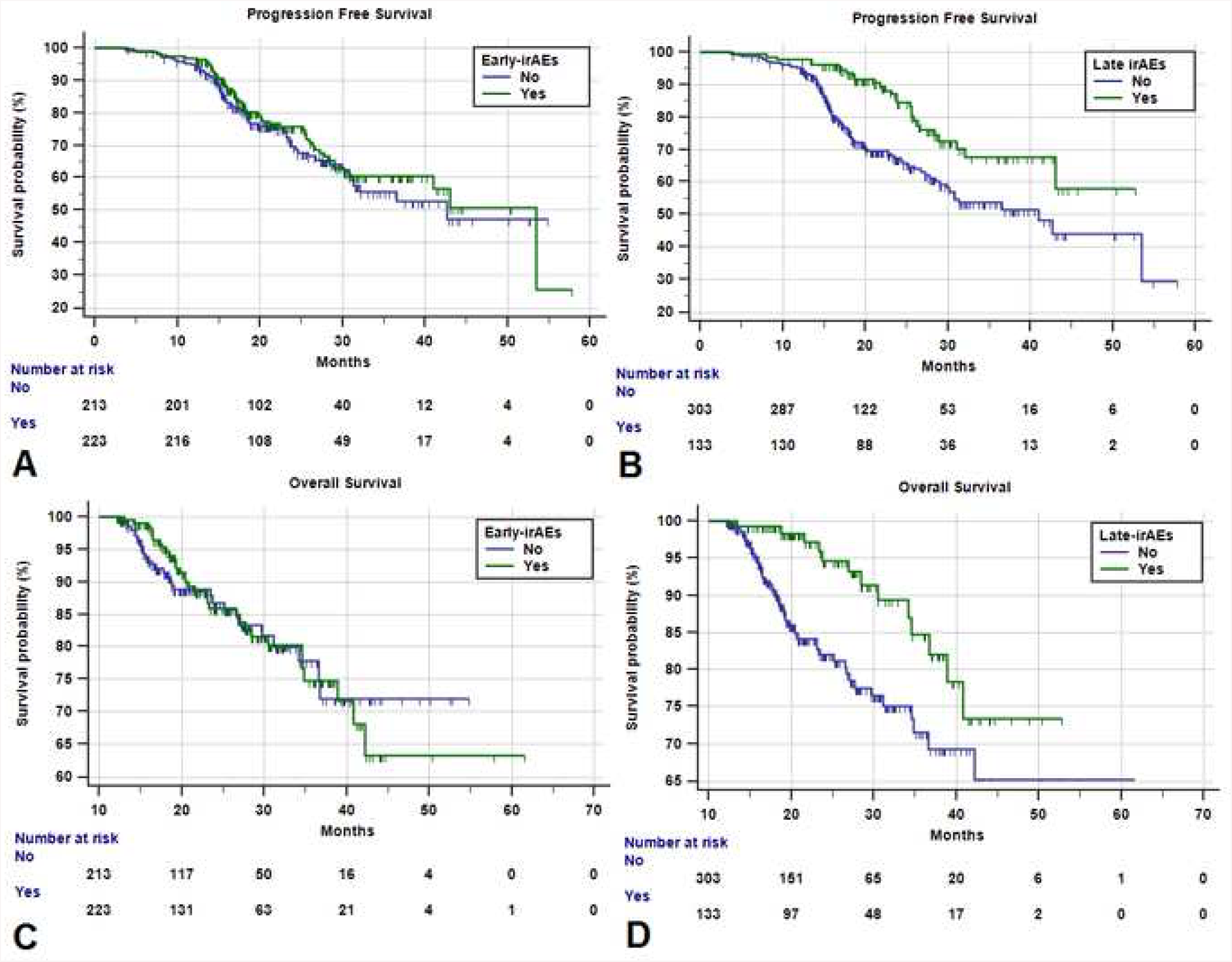
Kaplan-Meier survival curves. (A) Progression Free Survival according to the early-irAEs occurrence. (B) Progression Free Survival according to the late-irAEs occurrence. (C) Overall Survival according to the early-irAEs occurrence. (D) Overall Survival according to the late-irAEs occurrence.

Cumulative time-adjusted risk of disease progression according to both early- (HR=0.63 [95%CI: 0.30-1.29], p=0.204) and late-irAEs occurrence revealed no statistically significant differences (HR=0.75 [95%CI: 0.37-1.56], p=0.452) (Figure 2A-2C). Analogously, also time-adjusted cumulative risk of death according to both early-irAEs (HR=0.79 [95%CI: 0.34-1.86], p=0.598) and late-irAEs (HR=0.92 [95%CI: 0.49-1.74], p=0.811) did not show statistically significant differences (Figure 2B and 2D).

**Figure 2:**
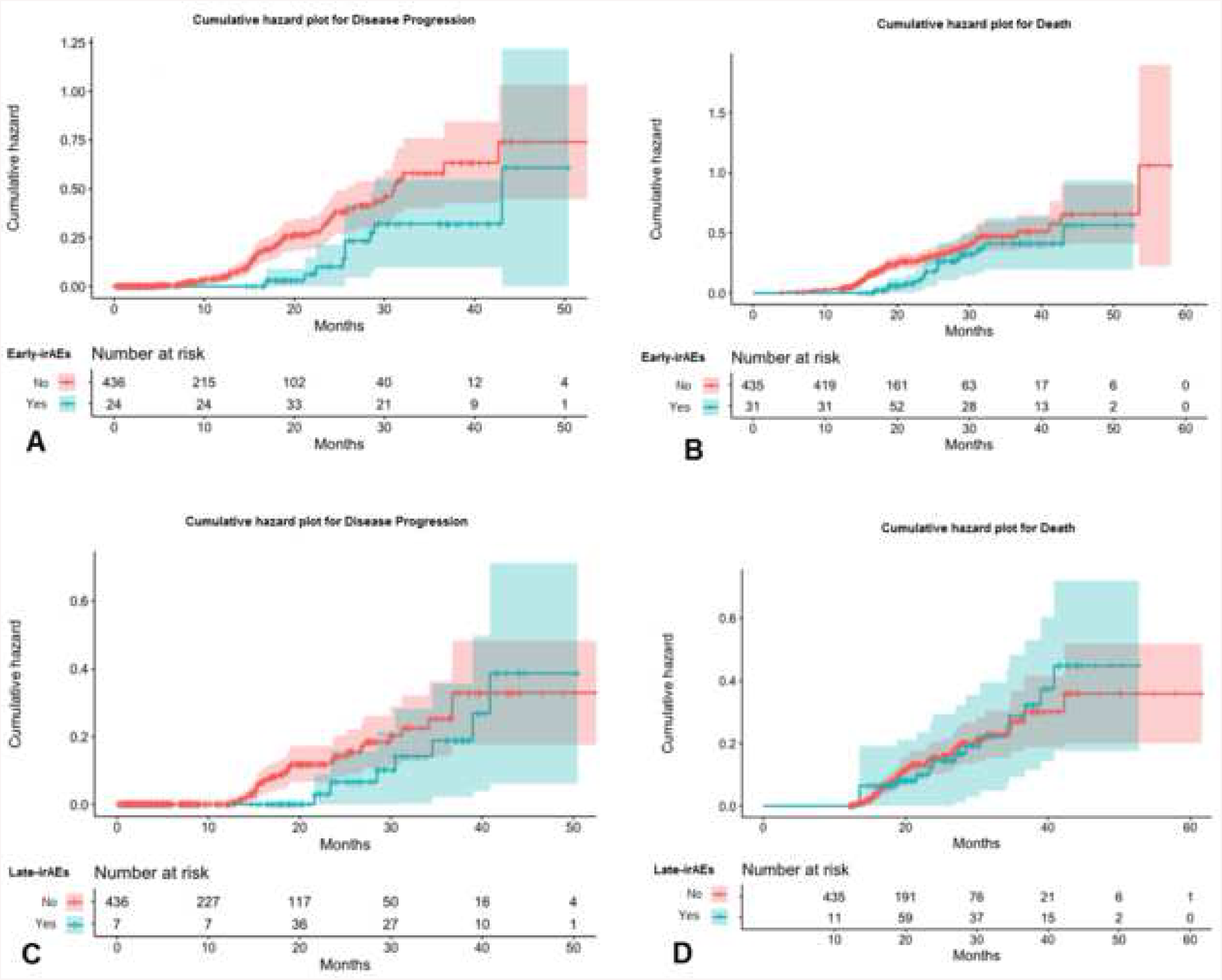
Cumulative hazard plots over time. (A) Disease progression according to the early-irAEs occurrence. (B) Death according to the early-irAEs occurrence. (C) Disease progression according to the late-irAEs occurrence. (D) Death according to the late-irAEs occurrence.

## Discussion

By definition, long-responders are positively selected patients; being able to be treated for a long time, with typically a lower tumor burden and fit to receive ICIs. In our study population, only 5.3% of patients had an ECOG-PS ≥2, while 34.4% had ≥3 involved organs. Positive selection might also have affected irAEs, both in terms of spectrum and grade. Considering any grade early-irAEs (51.1%) and G3 early-irAEs (4.8%), incidence was lower, compared to global incidence recently reported in clinical trials (any grade irAEs: 51.1%, G3/G4 irAEs: 4.8%) [22]. Moreover, no G4 irAEs were reported in the entire study population. Positive selection also explains the clinical management, with a relatively low need of systemic steroids administration (35.4%), and treatment interruption (15.2%), due to early-irAEs.

Globally, our study confirms the spectrum of all-grade irAEs described in both clinical trials and real-life studies with PD-1/PD-L1 inhibitors [22-24]; cutaneous (19.3%), endocrine (13.9%), and GI (10.8%) irAEs were the most commonly reported. Although they were not among the most frequent, rheumatologic irAEs had an incidence of 9.9%, higher than previously described in literature (0.7-5.1%) [24-25].

As irAEs could be considered a surrogate of effectiveness of ICIs [9, 10], positive selection might have also affected type and grade of irAEs, which not necessarily mirrors the spectrum of irAEs observed in general population of patients treated with ICIs. It has recently been reported that rheumatologic irAEs are more tissue-specific compared to non-rheumatologic, which might contribute to explain the higher incidence observed in our study [25]. A growing body of evidence suggests that checkpoint dysfunctions might directly contribute to autoimmune diseases pathogenesis and their clinical evolution, providing some insights regarding the underlying mechanisms of irAEs. A defective PD-1 inhibitory signalling might contribute to pathogenesis of rheumatoid arthritis [26] and giant cell arteritis [27]. Moreover, T-cell exhaustion can be related with a low severity of several autoimmune diseases [28]. ICIs treatment is associated to a shift from an exhausted T-cell phenotype to an active T-cell effector phenotype, and that might contribute to the pathogenesis of both rheumatic and non-rheumatic irAEs [25, 29-31].

Toxicity profile of cancer treatments could dramatically vary over time, leading to significant changes of risk/benefit ratio with exposure-adjusted safety analyses [32]. Our primary finding of a significantly lower incidence of late-irAEs compared to early-irAEs (30.3% vs 51.1%, p<0.0001), partially collide with that. Beyond positive selection, a little percentage of patients was still at risk of developing late-irAEs after the first 12 months of immunotherapy. This may partially explain how the cumulative incidence does not dramatically increase over time in clinical trials with PD-1/PD-L1 inhibitors [4-6]. Although it has been reported that irAEs have a certain tendency of recurrence after their resolution [33], in our study population only 22% of patients experienced a late recurrence of an early-irAEs. Even then, our results could be partly explained by positive selection, which probably excluded patients who experienced severe early-irAEs (which could have required permanent discontinuation ≤12 months, even after an attempt to rechallenge). Also, late-irAEs were easy to manage, requiring steroids administration (32.5%), temporary interruption (11.4%), or permanent immunotherapy discontinuation (11.4%) only in a minority of cases.

As for early-irAEs, cutaneous (11.5%), endocrine (4.6%) and GI (4.1%) were the most common irAEs, but significantly less frequent when compared to respective organ/system category early-irAEs (p=0.0062, p<0.0001, and p=0.0005, respectively). As for early-irAEs, also among late-irAEs, rheumatologic adverse events had a high crude incidence (6.7%). It has been reported that two markers of neutrophil activation, CD177 and CEACAM1, might be considered biomarkers for irAEs occurrence, particularly for GI toxicity [34]. Moreover, a recent study confirmed the possible role in irAEs pathogenesis, of overlapping antigens in tumor and healthy body sites [35]. These evidences suggest that irAEs are tissue-specific and tend to involve a specific system/organ. In accordance with that, it is already been reported that single-site are more frequent than multiple-site irAEs [36]. Interestingly, multiple-site late-irAEs (4.1%) were significantly less frequent compared to multiple-site early-irAEs (17%). These results allow us to speculate about the “tissue-specificity” of irAEs, which seems to be incremental, according to positive patient selection.

Several reports demonstrated a significant correlation between irAEs occurrence and ICIs effectiveness, even with land-mark analyses adjusted for exposure time [9, 10, 36, 37]. To date, the greatest criticism of this phenomenology has always been the immortal time bias, which implies that longer is the exposure time, the greater the risk of experience irAEs [11]. As a result, long responders would have a greater risk of developing irAEs overall. Recently, Eggermont et al. confirmed a strong association between irAEs and relapse free survival in patients with high-risk stage III melanoma, who were treated with 12-months adjuvant pembrolizumab, dealing with the time bias by using a time-dependent Cox model [21].

Clinical outcomes analysis according to the conventional Cox regression revealed that early-irAEs were not related with PFS and OS, while late-irAEs were significantly associated with improved PFS and OS. While positive selection of study population could surely explain that early-irAEs did not positively select patients for PFS and OS, the fact that all the patients (despite late-irAEs occurrence) have been treated for at least 12 months, might allows us to speculate that we overcame, at least partially, the lead-time bias. However, we must not fail in taking into account results according to the cumulative hazard estimation over time, which mitigate meaningfully the results. Cumulative time-adjusted risk of progression and death was not significantly reduced according to early- and late-irAEs occurrence indeed, despite the trends which were in accordance with the previous findings. Surely the immature survival data, with only 124 progressed patients (28.5%) and 63 deaths (14.5%) out of 436 patients, affected these results. Among study limitations we have to include also the retrospective design, which expose to selection biases, and the lack of centralized data review (imaging and toxicities).

## Conclusions

Although less frequent than early-irAEs, late-irAEs are quite common in long responders to PD-1/PD-L1 checkpoint inhibitors, and are different in terms of spectrum and grade. Despite late-irAEs occurrence seems to be related with improved PFS and OS. Time-adjusted analysis revealed that, in such a high selected population, cumulative risk of disease progression and death were not significantly reduced in patients who experienced late-irAEs. Prospective studies and a longer follow-up are warranted to confirm our results.

## Data Availability

the datasets used during the present study are available from the corresponding author upon reasonable request

## Ethics approval and consent to participate

All patients provided written, informed consent to treatment with immunotherapy. All patients alive at the time of data collection provided an informed consent for the present retrospective analysis. The procedures followed were in accordance with the precepts of Good Clinical Practice and the declaration of Helsinki. The study was approved by the respective local ethical committees on human experimentation of each institution, after previous approval by the coordinating center (Comitato Etico dell’Insubria, 126/2019).

## Authors’ contributions

All authors contributed to the publication according to the ICMJE guidelines for the authorship. All authors read and approved the submitted version of the manuscript (and any substantially modified version that involves the author’s contribution to the study). Each author have agreed both to be personally accountable for the author’s own contributions and to ensure that questions related to the accuracy or integrity of any part of the work, even ones in which the author was not personally involved, are appropriately investigated, resolved, and the resolution documented in the literature.

## Acknowledgements

This work was supported by the Consorzio Interuniversitario Nazionale per la Bio-Oncologia (CINBO).

## Consent for publication

Not applicable.

## Legend

**Supplementary file 1:** list of the oncological institutions of the study

## Conflict of Interest statement

**Please wait…**

If this message is not eventually replaced by the proper contents of the document, your PDF viewer may not be able to display this type of document.

You can upgrade to the latest version of Adobe Reader for Windows®, Mac, or Linux® by visiting http://www.adobe.com/go/reader_download.

For more assistance with Adobe Reader visit http://www.adobe.com/go/acrreader.

Windows is either a registered trademark or a trademark of Microsoft Corporation in the United States and/or other countries. Mac is a trademark of Apple Inc., registered in the United States and other countries. Linux is the registered trademark of Linus Torvalds in the U.S. and other countries.

